# CONSORT-TM: Text classification models for assessing the completeness of randomized controlled trial publications

**DOI:** 10.1101/2024.03.31.24305138

**Authors:** Lan Jiang, Mengfei Lan, Joe D. Menke, Colby J Vorland, Halil Kilicoglu

**Author notes:** Corresponding author: Halil Kilicoglu, PhD, School of Information Sciences University of Illinois Urbana-Champaign 614 E Daniel Street, Champaign, IL, 61820, USA.

## Abstract

**Objective:** To develop text classification models for determining whether the checklist items in the CONSORT reporting guidelines are reported in randomized controlled trial publications.

**Materials and Methods:** Using a corpus annotated at the sentence level with 37 fine-grained CONSORT items, we trained several sentence classification models (PubMedBERT fine-tuning, BioGPT fine-tuning, and in-context learning with GPT-4) and compared their performance. To address the problem of small training dataset, we used several data augmentation methods (EDA, UMLS-EDA, text generation and rephrasing with GPT-4) and assessed their impact on the fine-tuned PubMedBERT model. We also fine-tuned PubMedBERT models limited to checklist items associated with specific sections (e.g., Methods) to evaluate whether such models could improve performance compared to the single full model. We performed 5-fold cross-validation and report precision, recall, F1 score, and area under curve (AUC).

**Results:** Fine-tuned PubMedBERT model that takes as input the sentence and the surrounding sentence representations and uses section headers yielded the best overall performance (0.71 micro-F1, 0.64 macro-F1). Data augmentation had limited positive effect, UMLS-EDA yielding slightly better results than data augmentation using GPT-4. BioGPT fine-tuning and GPT-4 in-context learning exhibited suboptimal results. Methods-specific model yielded higher performance for methodology items, other section-specific models did not have significant impact.

**Conclusion:** Most CONSORT checklist items can be recognized reasonably well with the fine-tuned PubMedBERT model but there is room for improvement. Improved models can underpin the journal editorial workflows and CONSORT adherence checks and can help authors in improving the reporting quality and completeness of their manuscripts.

## BACKGROUND AND SIGNIFICANCE

Complete and transparent reporting in biomedical publications is critical for assessing the validity of research findings, promoting scientific integrity, and facilitating evidence-based decision-making in patient care and health policy^1–3^. However, poor reporting has been a persistent issue leading to potential biases and difficulties incorporating results into meta-analyses, complicating replication efforts, and ultimately undermining the trustworthiness of biomedical research^2,4,5^. Reporting guidelines have been developed to improve reporting for biomedical studies^5–9^ and provide the minimum list of information that must be reported in a publication. While they have been endorsed by many high-impact journals^10^, adherence to reporting guidelines remains inadequate^2,11,12^.

Randomized controlled trials (RCTs), when designed and conducted rigorously, remain the most robust method to determine the effectiveness of an intervention^13^. The CONSORT 2010 Statement^6,13^ is a reporting guideline for RCT results publications and consists of a checklist and participant flowchart. The checklist includes 25 items considered the minimum information needed to understand RCTs (e.g., outcomes, randomization, masking, harms). While CONSORT has been endorsed by many journals, publishers, and editorial organizations, systematic studies of current practices show poor reporting even in well-conducted RCTs^12–14^. Some studies have shown more complete reporting of CONSORT items over time^15^.

Adherence to CONSORT, and to reporting guidelines more broadly, remains low, partly because journal endorsement generally does not entail enforcement or verification. CONSORT implementation, where RCT submissions are scrutinized by journal editors for compliance before peer review, has been shown to improve reporting quality^16,17^. However, manual screening is labor-intensive and time-consuming for journal editors and staff. Automated screening, based on natural language processing (NLP) and machine learning approaches, could reduce the burden of manual checking, streamline the peer review process, and contribute to better reporting quality^18–20^.

In prior work, we developed a corpus of RCT result publications annotated with CONSORT checklist items (CONSORT-TM)^21^ and reported NLP models for recognizing the CONSORT items related to methodology (17 items)^15,21,22^. In this study, we extend our work by training and validating NLP models for the full CONSORT checklist at fine granularity (37 items). Our main contributions are as follows:

1. We develop and evaluate the first NLP models targeting automatic recognition of all CONSORT checklist items at fine granularity.
2. We compare different input representations and features for the task (context size, section information, sentence position).
3. We fine-tune a GPT-based model (BioGPT^23^) to study whether it confers any benefits over the models based on the BERT architecture^24^.
4. We assess in-context learning using GPT-4 for the task.
5. To address the data size and imbalance, we assess the ability of GPT-4 to generate useful training instances for this task and compare it to other data augmentation methods (EDA^25^, UMLS-EDA^26^).

## RELATED WORK

### Text classification in RCT articles

Most NLP research on RCT publications has focused on classifying sentences using the PICO framework (Population, Intervention, Comparator, Outcome) to aid the systematic review process and evidence-based medicine^27–34^. Other research has focused on automating risk of bias assessment using text classification^35–37^ and rhetorical classification of medical abstracts (e.g., Objective, Methods)^38–40^. Research on other key characteristics is less common, although methods have been reported for identifying study design^29,30,41^, sample size^28,37,41^, statistical methods^42^, and limitations^43^. The methods range from rule-based methods in early work^27,42^ to (semi-)supervised machine learning methods in later work^28,31,35,43^, including deep learning approaches of the recent years^32–34,38–41^.

We presented CONSORT-TM, a corpus which represents the most comprehensive annotation of RCT characteristics, to our knowledge^21^. We also trained NLP models for recognizing methodology items. A BioBERT-based model^44^ outperformed rule-based and traditional machine learning approaches. We used our best model to study reporting trends in more than 176K RCT publications published between 1966 and 2018, which showed an improvement in methodology reporting while also highlighting the shortcomings in the reporting of most items^15^.

### Generative large language models for biomedical literature mining

Generative large language models (LLMs) based on Transformer architecture^45^ (e.g., GPT family^46,47^, PaLM^48^, LLaMA^49^) have shown remarkable language generation capabilities and are increasingly applied to NLP tasks in the general domain^50^ and in the biomedical domain^51,52^. Domain-specific LLMs for the biomedical domain have also been trained (e.g., BioGPT^23^, Med-PaLM^51^). Prompt engineering for specific tasks has become an effective strategy for leveraging the in-context learning abilities of LLMs^52^. In the biomedical domain, fine-tuned BioGPT^23^ has shown superior performance to BERT-based models in document classification, while the performance of the GPT models in zero- or one-shot settings has been found to trail that of fine-tuned BERT models^53^. Most relevant to this work, a recent study used GPT-3.5 to check RCT reports on sports medicine and exercise science for adherence with 9 CONSORT checklist items, reporting accuracy in the range of 70-100%^54^. This study is limited in scope compared to ours and focuses on article-level binary decisions for the 9 items, not sentence classification.

## MATERIALS AND METHODS

### Dataset

The CONSORT-TM corpus^21^ consists of 50 RCT publications annotated at the sentence level with 37 CONSORT checklist items. It contains a total of 10,709 sentences, 4,845 (45%) of which are annotated with 5,246 labels (i.e., a multi-label corpus). Each article contains an average of 27.5 out of 37 fine-grained checklist items. In this work, we exclude the checklist item Background (2a) because it is too broad a category that virtually all papers report and checking its reporting was deemed unnecessary and focus on 36 items. We provide the CONSORT checklist items and their descriptions in Supplementary File Table S1.

### PubMedBERT fine-tuning

In prior work on CONSORT methodology reporting^21,22^, we fine-tuned the BioBERT model^44^. In more recent work^15^, we substituted the BioBERT model with PubMedBERT^55^, which has shown better performance in many biomedical NLP tasks. We continue to use PubMedBERT for multi-label sentence classification in this study. We excluded items 1a (whether the study is indicated as randomized in the title) and 1b (whether the abstract is structured) from the sentence classification model, because these items are article-level, unlike the rest of the items, and we use simple rules to recognize them (see below).

In previous work, we represented each input sentence as the concatenation of its enclosing section header and the sentence text. Here, we incorporate more contextual information into classification by taking into account the preceding and the following sentence, based on the observation that many CONSORT items are reported over several contiguous sentences (i.e., zones), and additional information from neighboring sentences could help in classification. The input representation is illustrated in Figure 1. It consists of three sentences (preceding, target, trailing) delimited by special [SEP] tokens and prepended by the [CLS] classification head, following earlier work^56,57^. Each sentence is prepended with the list of (nested) section headers associated with the sentence (e.g., *Methods Patients* [SENTENCE]). The [CLS] token representation generated by PubMedBERT encoder is fed into a fully connected layer and the sigmoid function is used for multi-label classification.

**Figure 1.**
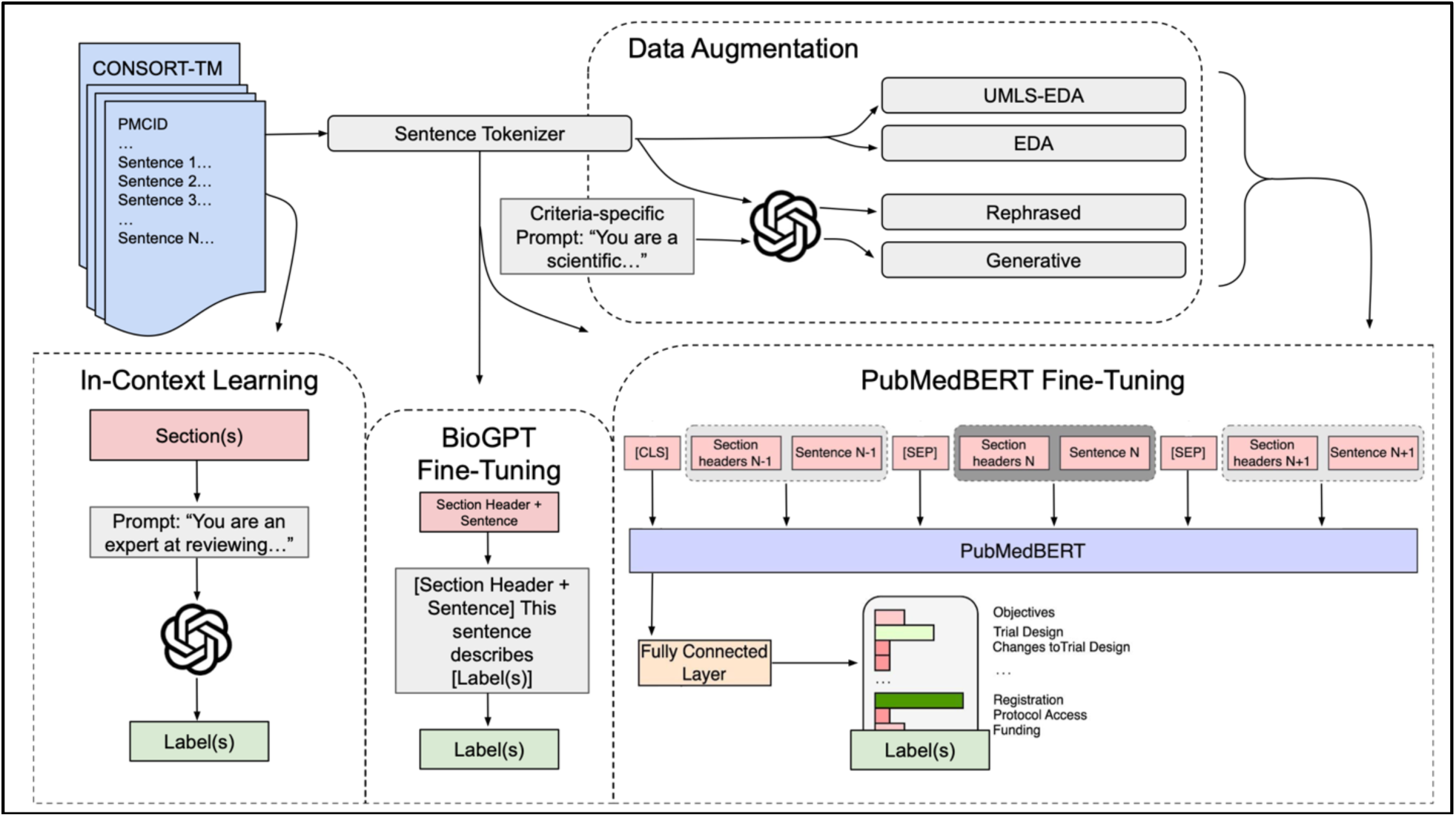
Experimental flow including PubMedBERT fine-tuning, data augmentation strategies, few-shot prompting, and BioGPT fine-tuning. The PubMedBERT model takes sentences surrounding the target sentence into consideration. The prediction is made on the [CLS] token representation.

We also leveraged sentence position in the article as an additional feature, based on the observation that checklist items tend to be discussed in a predictable order in an article. For example, the first few sentences of the Methods section often discuss item 3a (Trial Design). We concatenated sentence position embedding with the sentence representation to incorporate this information. We experimented with absolute and relative positions. We used an embedding layer to encode absolute position. We convert relative position (continuous value) into a categorical value by creating 10 bins (0-0.1, 0.1-0.2, etc.) first and then used an embedding layer for encoding it.

CONSORT checklist is organized by sections in which the items are expected to be reported. We examined whether models trained on specific sections and items specific to those sections could lead to better performance than a single model trained on full articles and all labels. For these experiments, we created models specific to Methods, Results, and Discussion sections. We did not create an Introduction-specific model, because we consider only one relevant label in this section (2b (Objectives)).

### Data augmentation

CONSORT-TM is relatively small and some checklist items are infrequently reported (e.g., 6b (Changes to Outcomes)). This has led to poor performance for rare items in previous work^21,22^. In this study, we leveraged the text generation capabilities of LLMs to improve the quality of data augmentation and compared it to simpler approaches we explored in previous work (EDA^24^, UMLS-EDA^25^). Our goal was to assess whether this type of data augmentation could improve model performance for rare labels and enhance generalizability.

#### Prompt-based augmentation using GPT-4

GPT-4 can generate fluent, human-like text, even about complex topics like medicine^58^. Previous work has demonstrated that GPT-based models can be used for data augmentation to improve a model’s performance; for example, AugGPT reports a framework that uses ChatGPT to rephrase existing text instances^59s^.

We adapted AugGPT^59^ to paraphrase the instances of rare labels in our corpus. In addition, we used GPT-4 to generate completely new sentences based on label descriptions, referred to here as generative instances. For paraphrased instances, we used the labels of the original instance. For generative instances, we applied the label of the criterion description used within the prompt.

We augmented data for categories that have fewer than 100 samples in the entire corpus. These categories are: 3b (Changes to Trial Design), 6b (Changes to Outcomes), 7b (Interim Analyses and Stopping Guidelines), 9 (Allocation Concealment Mechanism), 11b (Similarity of Interventions), 12b (Statistical Methods for Other Analyses), 14b (Trial Stopping) and 21 (Generalizability). To better isolate the effect of the different augmentation strategies, we evaluated the performance on a version of the model architecture that only uses the target sentence for classification, as well as on our best-performing architecture, which uses section headings and surrounding sentences. We used the prompts shown in Table 1 to generate sentences using the OpenAI GPT-4 API (generation performed on Sept. 11, 2023). In both prompts, N indicates the number of instances to generate. Each instance consists of a preceding sentence, a positively labeled sentence, and a trailing sentence to be used by the model.

**Table 1.** GPT-4 prompts used for data augmentation.

|  |  |
| --- | --- |
| Rephrasing | *You are a scientific researcher writing a research paper for randomized clinical trials. Please rephrase the following sentence N times: [POSITIVELY LABELED SENTENCE]. For the N rephrased sentences, also provide an example preceding sentence and an example trailing sentence. Vary the sentence structure and sentence complexity for each sentence. Also, vary the use of introductory prepositional phrases in all sentences.* |
| Generative | *You are a scientific researcher writing a research paper for randomized clinical trials. From your research paper, please provide N different 1 sentence examples of the following: [EDITED CHECKLIST ITEM DESCRIPTION, WITH SPECIFICS]. For each example, also provide an example preceding sentence and an example trailing sentence. Vary the sentence structure and sentence complexity for each example. Also, vary the use of introductory prepositional phrases in all examples.* |

For rephrased instances, N was set to 6, following AugGPT. For generative instances, we iteratively prompted GPT-4 setting N to 6 or 8 until we accumulated 100 total instances (13 times).

Item descriptions used for the generative prompt were adapted from Moher et al.^13^. For all descriptions, the phrase “with reasons” was changed to “with specifics”, and irrelevant text (e.g., “when applicable” or “if relevant”) and examples (e.g., “such as eligibility criteria”) were removed to improve the overall quality and diversity of GPT-4 responses.

#### Easy data augmentation (EDA)

As a simpler alternative to prompt-based data augmentation, we also generated examples using EDA^24^ and its variant, UMLS-EDA^25^. EDA^24^ is a rule-based method that synthesizes samples via simple modifications to the original sentence, including random deletion, random insertion, random swap, and synonym replacement based on WordNet. UMLS-EDA^25^ is an adaptation of EDA that additionally uses synonym replacement based on the UMLS^60^. For both methods, we generated six variations for each original sample, for consistency with the rephrasing approach. We only used the instances with a single label in the corpus for data augmentation.

### In-context learning with GPT-4

To assess the few-shot learning ability of GPT-4 for our task, we prompt GPT-4 to directly infer whether a sentence in the article reports a specific CONSORT checklist item. The prompt given to GPT-4 consists of the task description, checklist item descriptions, examples (in one-shot and five-shot settings), and the entire article. We provide the full prompt in Supplementary File.

### BioGPT fine-tuning

We also fine-tuned BioGPT^23^ for the task, formulating the task as text generation. BioGPT is based on GPT-2^46^ and trained using biomedical literature and has shown improved performance on text classification tasks over BERT-based architectures^23^. We used a fine-tuning formulation similar to that proposed in Luo et al.^23^ for document classification. We generate the sequence using the format “[SENTENCE]. *This sentence describes* [LABEL]”. [SENTENCE] includes the section header information. We also learned one virtual token to better steer the language model to our task. We let BioGPT complete the sentence to perform the sentence classification task.

### Article-level classification for items 1a and 1b

CONSORT checklist items related to title and abstract (1a and 1b) are document-level. We developed a simple rule-based approach for these items. For 1a (is the study described as “randomized” in the title?), we check the stems in the title for the presence of *random*, *randomis*, and *randomiz*. For 1b (is the abstract structured?), we check whether the abstract starts with a structured abstract header included in the list developed by the National Library of Medicine^61^.

### Evaluation

To evaluate the fine-tuned PubMedBERT and BioGPT models, we used group 5-fold cross-validation, ensuring that sentences in one article can only be in training or test set for each cross-validation run. We provide the experimental settings in Supplementary File.

To evaluate in-context learning with GPT-4, we randomly sampled 10 articles as the test set, because the model yielded only modest performance in preliminary experiments (see Results) and using OpenAI API for GPT-4 incurs significant cost.

Following previous work, we used precision, recall, and their harmonic mean, F1 score, as the main evaluation criteria. We report micro- and macro-averaged results and the area under the ROC curve (AUC). To observe whether different input representations and data augmentation approaches led to statistically significant differences in model performances compared to the baseline, we used McNemar’s test^62^, adopting the approach outlined by Gillick and Cox^63^. In addition to sentence-level results, we also report article-level results, i.e., whether the model correctly predicts if the article includes at least one sentence relevant to the checklist item. This is a more lenient evaluation, which can be appropriate in some use cases, such as guideline adherence checks or large-scale reporting analyses^15^.

## RESULTS

### High-level comparison of PubMedBERT and GPT-based models

Table 2 shows the performance of the sentence classification models. The results show that section headers contribute significantly to PubMedBERT performance. Prepending all relevant section headers outperforms prepending only the innermost or outermost section header. Incorporating positional information does not improve results. On the other hand, incorporating context from surrounding sentences yields the best performance, specifically by improving recall. We consider this model (PubMedBERT using surrounding context, prepending section headers to sentences, and using [CLS] token representation) as our main model.

**Table 2.** Overall model performance for CONSORT sentence classification over 5-fold cross-validation. PubMedBERT results with different input representations and additional features are shown. The best performances are marked in bold. Standard deviation is shown in parentheses. We do not calculate AUC scores for BioGPT and GPT-4 because predicted probabilities are not available. * indicates that the performance difference with the baseline PubMedBERT model (A), calculated using McNemar’s test, is statistically significant (*p* < .0001). AUC: Area under ROC curve.

| Model | Precision | Recall | F<sub>1</sub> | Macro-F<sub>1</sub> | AUC |
| --- | --- | --- | --- | --- | --- |
| Baseline PubMedBERT [A] | 0.66 (0.03) | 0.64 ( 0.04) | 0.65 (0.03) | 0.59 (0.02) | 0.94(0.01) |
| (A) + innermost section headers prepended * | 0.69 (0.03) | 0.66 (0.04) | 0.68 (0.03) | 0.60 (0.02) | 0.95 (0.01) |
| (A) + outermost section headers prepended* | 0.70 (0.04) | 0.67 (0.04) | 0.69 (0.04) | 0.62 (0.01) | 0.95 (0.01) |
| (A) + all section headers prepended [B] * | 0.71 (0.02) | 0.69 (0.04) | 0.70 (0.03) | 0.63 (0.03) | **0.96 (0.00)** |
| (B) + absolute sentence position * | **0.72 (0.03)** | 0.69 (0.03) | **0.71 (0.03)** | 0.62 (0.03) | 0.95 (0.00) |
| (B) + relative sentence position * | 0.71 (0.02) | 0.69 0.03) | 0.70 (0.02) | 0.63 (0.01) | 0.95 (0.01) |
| (B) + contextual information * | **0.72 (0.02)** | **0.71 (0.03)** | **0.71 (0.02)** | **0.64 (0.02)** | **0.96 (0.01)** |
| BioGPT * | 0.68 | 0.68 | 0.68 | 0.55 | - |
| GPT-4 (zero-shot) | 0.48 | 0.54 | 0.51 | 0.49 | - |
| GPT-4 (one-shot) | 0.50 | 0.45 | 0.48 | 0.48 | - |
| GPT-4 (five-shot) | 0.50 | 0.42 | 0.46 | 0.47 | - |

BioGPT fine-tuning yields modest improvement over the baseline PubMedBERT model; however, it is outperformed by PubMedBERT models which use richer input representations. Zero-shot in-context learning with GPT-4, however, yields poorer performance compared to fine-tuned models. Surprisingly, providing example sentences (one-shot and five-shot) degraded the GPT-4 performance further. GPT-4 showed improved recognition of some rare items, such as 7b (Interim Analysis/Stopping Guidelines), 9 (Allocation Concealment), and 11b (Similarity of Interventions); while its performance on common items, such as 6a (Outcomes), and 12a (Statistical Methods for Outcomes) was notably lower. Item-level results for the models are shown in Supplementary File Tables S2-4.

### Item-level results for the best-performing PubMedBERT model

The best-performing PubMedBERT model yields over 0.8 F1 score for 8 items at the sentence level (out of 34), all of which contain more than 100 instances in the dataset (Supplementary File Table S2). The model performance remains relatively low in classifying infrequently reported items. F1 score remains under 0.5 for another 9 items, most of which have fewer than 100 instances in the dataset (3b, 6b, 7b, 9, 12b, and 21). Some CONSORT items are multi-part (indicated by a and b in the item numbers) and in some cases the model struggles with distinguishing these closely related items. For example, item 12b (Statistical Methods for Other Analyses) is often confused with 12a (Statistical Methods for Outcomes). The performance is highest for items related to Introduction (0.89 F1 for 2b (Objectives)), followed by Methods sections (0.75 F1). It is lowest for Results-related items (0.62 F1). The performance on items not associated with specific sections (items 23-25) is over 0.8 F1.

The article-level classification results combine both the best-performing PubMedBERT model (2b to 25) and the rule-based method (1a and 1b), reaching high micro-F1 score of 0.95 (Supplementary File Table S2). 26 out of 36 items are recognized with F1 score of 0.8 or higher, while the verification of infrequent items is still challenging. Both items 1a and 1b are recognized accurately, showing that the simple rules are sufficient for these items.

### Data augmentation

We present samples generated via data augmentation in Supplementary File Table S5. GPT-4 is able to generate to coherent sentences in “generative” setting from the item descriptions. Using GPT-4 for rephrasing also seem to largely preserve the semantic content of the sentence. On the other hand, EDA and UMLS-EDA methods do not preserve meaning.

To assess the contribution of data augmentation, we use both the baseline PubMedBERT model and the best-performing model. The results are presented in Table 3. We observe that data augmentation does not improve results of the best-performing model. For the baseline model (sentence text only), UMLS-EDA improves the results most (2 percentage points). A closer analysis reveals that different methods improve the performance of infrequent items (reflected by the increase in macro-F1), while this improvement is often offset by performance reduction in more common items.

**Table 3.** Performance of CONSORT sentence classification with different data augmentation methods. The average of micro-precision, recall, and F1, as well as macro F1 over 5-fold cross-validation are reported. The performance differences between the data augmentation method and the original models are not statistically significant. Standard deviation and AUC are not shown for brevity.

| Input | Baseline PubMedBERT |  |  |  | Best-performing model |  |  |  |
| --- | --- | --- | --- | --- | --- | --- | --- | --- |
|  | *Prec.* | *Recall* | *F<sub>1</sub>* | *Macro-F<sub>1</sub>* | *Prec.* | *Recall* | *F<sub>1</sub>* | *Macro-F<sub>1</sub>* |
| Original | 0.65 | 0.64 | 0.64 | 0.59 | 0.72 | 0.71 | 0.71 | 0.64 |
| + GPT-4 generative | 0.65 | 0.63 | 0.64 | 0.59 | 0.71 | 0.71 | 0.71 | 0.63 |
| + GPT-4 rephrasing | 0.66 | 0.64 | 0.65 | 0.59 | 0.72 | 0.70 | 0.71 | 0.62 |
| + EDA | 0.66 | 0.64 | 0.65 | 0.59 | 0.73 | 0.70 | 0.71 | 0.64 |
| + UMLS-EDA | 0.67 | 0.65 | 0.66 | 0.60 | 0.72 | 0.71 | 0.71 | 0.66 |

### Comparison with section-specific models

The comparison of the model trained on full articles and label set with the models trained on specific sections and related labels are shown in Table 4. Training a Methods-specific model using the Methods sentences yielded better micro-F1 score than the single model trained on the full article. This finding held for Results and Discussion sections, albeit to a smaller degree. The effect of section-specific training seems to be to improve precision with some recall loss. Macro-F1 scores were higher with the single model, suggesting that section-specific models primarily improve the performance of the common items.

**Table 4.** Performance of sentence classification models trained on specific sections or on the entire article. The average micro-precision, recall, F1, as well as macro-F1 scores over 5-fold cross-validation are reported. Standard deviation and AUC are not shown for brevity.

| Section | Trained on specific section/label set |  |  |  | Trained on full articles/label set |  |  |  |
| --- | --- | --- | --- | --- | --- | --- | --- | --- |
|  | *Prec.* | *Recall* | *F<sub>1</sub>* | *Macro-F<sub>1</sub>* | *Prec.* | *Recall* | *F<sub>1</sub>* | *Macro-F<sub>1</sub>* |
| Methods | 0.79 | 0.78 | 0.79 | 0.61 | 0.71 | 0.78 | 0.75 | 0.63 |
| Results | 0.64 | 0.62 | 0.63 | 0.59 | 0.59 | 0.65 | 0.62 | 0.64 |
| Discussion | 0.70 | 0.67 | 0.68 | 0.57 | 0.64 | 0.68 | 0.67 | 0.60 |

## DISSCUSSION

This study is the first to present an automated approach for recognizing all CONSORT checklist items in RCT results publications. The overall performance of the best model is reasonable (0.71 micro-F1 with balanced precision and recall). For common items such as Eligibility Criteria (4a), Outcomes (6a), Sample Size Determination (7a), and Registration (23), its performance is over 0.8 F1 score, indicating that the model could be used for recognizing such items in practice. The performance is lowest on rare items, such as Changes to Trial Design (3b), Changes to Outcomes (6b), and Allocation Concealment (9). Recognition of CONSORT items at the article level is high (0.95 micro-F1). While article-level predictions may be less useful than sentence-level predictions, they can facilitate automatic screening by pointing out whether or not a publications reports a specific item.

The best-performing classifier is a fine-tuned PubMedBERT model that uses as input the target sentence as well as the surrounding sentences, each prepended with their section headers. This indicates the utility of longer context and document structure for the task. This is not surprising, given that some CONSORT items are reported over passages and the section header are sometimes directly related to the CONSORT item (e.g., *Primary outcomes*). The impact of incorporating longer context versus document structure is similar and they act synergistically to further improve the performance, although this additional improvement is small. We leave the investigation of whether even longer contexts could lead to further performance improvement to future work. An analysis of the errors made by the best-performing PubMedBERT model is presented in Supplementary File.

### Generative models

The generative models for sentence classification (BioGPT fine-tuning and zero- or few-shot in-context learning with GPT-4) underperformed the best PubMedBERT model by significant margins. Similar to PubMedBERT models, BioGPT did well for some common items (e.g., 7a (Sample Size Determination), 0.87 F1), while its performance was poor for rare items and some multi-part items (Supplementary File Table S3). BioGPT fine-tuning involved only the target sentence, and adding surrounding sentence could possibly improve performance; however, fine-tuning BioGPT is much more computationally intensive to fine-tuning PubMedBERT. BioGPT is based on GPT-2^46^, and using more recent domain-specific models such as PMC-LLaMA^64^ could be a more promising avenue.

In-context learning with GPT-4 failed to achieve satisfactory results, even for common items. Surprisingly, providing examples (one- or few-shot) did not improve upon zero-shot setting. Existing studies point out that GPT models are sensitive to the prompts and even the order of elements in the prompts; therefore, it may be possible to design better prompts to enhance in-context learning. We randomly sampled demonstration examples for one- or few-shot settings; selecting examples similar to the target sentence could improve results. At the same time, our results with GPT-4 are consistent with other comparisons of GPT-4 in-context learning with fine-tuned models for text classification^53^. A more comprehensive study of prompting strategies for the task is needed in the future.

### Data augmentation

The effect of data augmentation on PubMedBERT fine-tuning was minimal, which is consistent with our previous findings^22^. To our surprise, GPT-4 based approaches underperformed EDA-based approaches, UMLS-EDA in particular, even though they produced more meaningful, generally semantically coherent sentences. Our findings with GPT-4 contrast other studies that found that synthetic data generation with LLMs led to improved performance of downstream tasks^65^. In GPT-4-based augmentation, we only provide the target sentence for rephrasing and let GPT-4 generate corresponding preceding and trailing sentences. This may have led to inconsistencies between the sentences generated and reduced the effectiveness of this approach. Data augmentation had a more pronounced effect when it is used to enhance the baseline PubMedBERT model, in contrast to the best-performing model that uses longer contexts. This suggests that longer contexts, to some extent, could compensate for data scarcity.

### Section-specific training

Our comparison of section-specific model training with training of a single CONSORT model was inconclusive. Methods-specific model worked better on methodology items than the more comprehensive model. On the other hand, the results for Results and Discussion sections were mostly similar. Precision based on section-specific training was notably higher, which may be desirable in some cases. However, because the differences are minor, it seems more efficient to train and perform predictions using a single full model.

### Limitations

A limitation of our study is the limited training data size, especially for the infrequently reported CONSORT checklist items. We attempted to address this issue using data augmentation; however, the effect was minimal. Given the scarcity of data, it might be reasonable to resort to rule-based methods for some rare items, such as 3b (Changes to Trial Design). At the same time, it is necessary to develop larger datasets, which is challenging, as it requires significant domain expertise. Distant supervision approaches leveraging unlabeled data from the literature could be a promising avenue. Our exploration of generative models was limited. A more systematic exploration of prompting strategies is needed in future work.

We have focused on recognizing sentences reporting CONSORT checklist items, a first step toward assessing adherence (e.g., whether the statistical methods used are appropriate), which we have not attempted in this study. We leave this much more challenging task for future work.

## CONCLUSIONS

In this study, we extended our earlier work to recognize all CONSORT checklist items in RCT publications. A PubMedBERT fine-tuned model using surrounding contexts and article structure yielded the best performance. We did not observe significant benefits from using LLMs for data augmentation or in-context learning, or fine-tuning them. We also did not observe an advantage of training section-specific models.

In future work, we aim to improve the models further for practical use. We plan to achieve this by extending the annotated corpus and the models, and making the models more efficient by employing techniques such as distillation. With further enhancements, the models could assist journals in checking for CONSORT compliance in a human-in-the-loop setting and help the authors in improving the completeness and transparency of their manuscripts. We plan to extend our models to assess the extent to which articles are CONSORT-compliant, potentially increasing the practical utility of the models.

## DATA AVAILABILITY

CONSORT-TM dataset, the PubMedBERT models, and source code are available at https://github.com/ScienceNLP-Lab/RCT-Transparency.

## ACKNOWLEDGMENTS

This work was supported by the National Library of Medicine of the National Institutes of Health under the award number R01LM014079. The content is solely the responsibility of the authors and does not necessarily represent the official views of the National Institutes of Health. The funder had no role in considering the study design or in the collection, analysis, interpretation of data, writing of the report, or decision to submit the article for publication.

## COMPETING INTERESTS STATEMENT

The authors state that they have no competing interests to declare.

## CONTRIBUTORSHIP STATEMENT

**LJ:** Conceptualization, Methodology, Software, Validation, Formal analysis, Investigation, Writing – Original draft, Writing – Review & Editing. **ML:** Methodology, Software, Validation, Formal analysis, Investigation, Writing – Original draft, Writing – Review & Editing. **JM:** Methodology, Software, Validation, Formal analysis, Investigation, Writing – Original draft, Writing – Review & Editing. **CJV:** Methodology, Software, Formal analysis, Writing – Review & Editing. **HK:** Conceptualization, Methodology, Data curation, Investigation, Supervision, Project administration, Funding acquisition, Writing – Original draft, Writing – Review & Editing.

## Notes

### Competing Interest Statement

The authors have declared no competing interest.

### Funding Statement

This study was funded by the National Library of Medicine of the National Institutes of Health under the award number R01LM014079. The content is solely the responsibility of the authors and does not necessarily represent the official views of the National Institutes of Health. The funder had no role in considering the study design or in the collection, analysis, interpretation of data, writing of the report, or decision to submit the article for publication.

