## Supplementary File for "CONSORT-TM: Text classification models for assessing the completeness of randomized controlled trial publications"

### 1. CONSORT checklist

The CONSORT checklist items and their descriptions are provided below in Table S1. We refer the reader to Moher et al.[1] for reporting examples of each item.

**Table S1.** CONSORT checklist items, corresponding item numbers, and the sections to which they typically belong.

| Section | Item | Item No | Description |
|---|---|---|---|
| Title | | 1a | Identification as a randomized trial in the title |
| Abstract | | 1b | Structured summary of trial design, methods, results, and conclusions |
| Introduction | Background | 2a | Scientific background and explanation of rationale |
| | Objectives | 2b | Specific objectives or hypotheses |
| Methods | Trial design | 3a | Description of trial design (such as parallel, factorial) including allocation ratio |
| | | 3b | Important changes to methods after trial commencement (such as eligibility criteria), with reasons |
| | Participants | 4a | Eligibility criteria for participants |
| | | 4b | Settings and locations where the data were collected |
| | Interventions | 5 | Interventions for each group with sufficient details to allow replication, including how and when they were administered |
| | Outcomes | 6a | Completely defined pre-specified primary and secondary outcome measures, including how and when they were assessed |

| | | | |
|---|---|---|---|
| | | 6b | Any changes to trial outcomes after the trial commenced, with reasons |
| | Sample size | 7a | How sample size was determined |
| | | 7b | When applicable, explanation of any interim analyses and stopping guidelines |
| | Randomization: Sequence generation | 8a | Method used to generate the random allocation sequence |
| | | 8b | Type of randomization: details of any restriction (such as blocking and block size) |
| | Randomization: Allocation concealment | 9 | Mechanism used to implement the random allocation sequence (such as sequentially numbered containers), describing any steps taken to conceal the sequence until interventions were assigned |
| | Randomization: Implementation | 10 | Who generated the random allocation sequence, who enrolled participants, and who assigned participants to interventions |
| | Blinding | 11a | If done, who was blinded after assignment to interventions (for example, participants, care providers, those assessing outcomes) and how |
| | | 11b | If relevant, description of the similarity of interventions |
| | Statistical methods | 12a | Methods used to compare groups for primary and secondary outcomes |
| | | 12b | Methods for additional analyses, such as subgroup analyses and adjusted analyses |
| Results | Participant flow | 13a | For each group, the number of participants who were randomly assigned, received intended treatment, and were analyzed for the primary outcome |
| | | 13b | For each group, losses and exclusions after randomization, together with reasons |
| | Recruitment | 14a | Dates defining the periods of recruitment and follow-up |
| | | 14b | Why the trial ended or was stopped |

| | | | |
|---|---|---|---|
| | Baseline data | 15 | A table showing baseline demographic and clinical characteristics for each group |
| | Numbers analyzed | 16 | For each group, number of participants included in each analysis and whether the analysis was by original assigned groups |
| | Outcomes and estimation | 17a | For each primary and secondary outcome, results for each group, and the estimated effect size and its precision (such as 95% confidence interval) |
| | | 17b | For binary outcomes, presentation of both absolute and relative effect sizes |
| | Ancillary analyses | 18 | Results of any other analyses performed, including subgroup analyses and adjusted analyses, distinguishing pre-specified from exploratory |
| | Harms | 19 | All important harms and unintended effects in each group |
| Discussion | Limitations | 20 | Trial limitations, addressing sources of potential bias, imprecision, and if relevant, multiplicity of analyses |
| | Generalizability | 21 | Generalizability (external validity, applicability) of the trial findings |
| | Interpretation | 22 | Interpretation consistent with results, balancing benefits and harms, and considering other relevant evidence |
| Other | Registration | 23 | Registration number and name of trial registry |
| | Protocol | 24 | Where the full trial protocol can be accessed, if available |
| | Funding | 25 | Sources of funding and other support (such as supply of drugs), role of funders |

### 2. Prompt for GPT-4 in-context learning

The GPT-4 prompt for in-context learning consists of the following ordered elements:

- **Task:** A short paragraph that explains the task, describes the input format, and constrains the response format. The prompt for this part is as follows:

*"You are an expert at reviewing scientific articles for transparency. I am providing you a CONSORT checklist table, and a list of lists of the sentence id number, section header, and sentence from the article. Based on that CONSORT table, only return a Python list of lists (no additional text, and no html returns needed) with the sentence id number and which CONSORT Item No you would match it to (as a string). Be strict with your matches. Some sentences may have multiple labels (if so, separate them by a comma), and some may have none (in those cases, make it a ['0']).*

- **Guidelines:** The CONSORT checklist, including related section, item number, and detailed description of each item.
- **Examples:** We experiment with zero-shot, one-shot and five-shot settings. In zero-shot setting, no examples are provided; in-context learning only relies on task definition and guidelines. In one-shot setting, we pick one sentence randomly from the samples in CONSORT-TM for each label. In five-shot setting, we randomly select five examples.
- **Article:** We add the entire article after the task definition, guideline description, and examples.

### 3. Experimental settings

We used HuggingFace implementation of the PubMedBERT (*BiomedNLP-PubMedBERT-base-uncased-abstract-fulltext*) model. We use the following hyperparameters: batch size (4), learning rate (1e-5), and dropout rate of 0.1. The learning rate of the fully connected layer is 1e-3. In each experimental run, we trained the model for 20 epochs.

We fine-tuned BioGPT which is pre-trained on PubMed abstracts from scratch for 20K steps with a peak learning rate of 1e-5 and 1000 warm-up steps. For GPT-4, we set the temperature to 1 when performing data augmentation to increase the creativity of the responses and 0 when performing direct inference to ensure that the responses are consistent.

### 4. Item-level results

**Table S2:** Sentence-level and article-level performance of the best-performing PubMedBERT model (2b-25) and the rule-based method (1a and 1b). The mean and standard deviation of precision, recall, micro-$F_1$, and macro-F1 scores over five folds in cross-validation are reported. Standard deviation is shown in parentheses.

| **CONSORT item** | **Sentence-Level** | | | **Article-Level** |
|---|---|---|---|---|
| | **Precision** | **Recall** | **$F_1$** | **$F_1$** |
| Title Randomized (1a) | - | - | - | 1.00 |
| Structured Abstract (1b) | - | - | - | 1.00 |
| Objectives (2b) | 0.90 (0.09) | 0.88 (0.05) | 0.89 (0.06) | 0.96 (0.05) |
| Trial Design (3a) | 0.82 (0.13) | 0.69 (0.11) | 0.73 (0.05) | 0.83 (0.07) |
| Changes to Trial Design (3b) | 0.00 (0.00) | 0.00 (0.00) | 0.00 (0.00) | 0.00 (0.00) |
| Eligibility Criteria (4a) | 0.92 (0.09) | 0.86 (0.06) | 0.88 (0.02) | 0.99 (0.02) |
| Data Collection Setting (4b) | 0.75 (0.08) | 0.72 (0.14) | 0.72 (0.05) | 0.57 (0.33) |
| Interventions (5) | 0.75 (0.06) | 0.74 (0.07) | 0.74 (0.05) | 0.97 (0.05) |
| Outcomes (6a) | 0.83 (0.05) | 0.86 (0.09) | 0.84 (0.07) | 1.00 (0.00) |
| Changes to Outcomes (6b) | 0.00 (0.00) | 0.00 (0.00) | 0.00 (0.00) | 0.00 (0.00) |
| Sample Size Determination (7a) | 0.91 (0.08) | 0.88 (0.09) | 0.89 (0.07) | 1.00 (0.00) |
| Interim Analyses/ Stopping Guidelines (7b) | 0.65 (0.38) | 0.43 (0.36) | 0.48 (0.31) | 0.59 (0.34) |
| Sequence Generation (8a) | 0.81 (0.15) | 0.72 (0.20) | 0.76 (0.17) | 0.60 (0.18) |
| Randomization Type (8b) | 0.79 (0.14) | 0.74 (0.13) | 0.76 (0.10) | 0.80 (0.12) |
| Allocation Concealment (9) | 0.59 (0.39) | 0.35 (0.15) | 0.40 (0.19) | 0.66 (0.42) |
| Randomization Implementation (10) | 0.69 (0.23) | 0.59 (0.17) | 0.63 (0.17) | 0.83 (0.11) |
| Blinding (11a) | 0.77 (0.13) | 0.64 (0.17) | 0.70 (0.15) | 0.89 (0.05) |
| Similarity of Interventions (11b) | 0.55 (0.27) | 0.49 (0.33) | 0.51 (0.31) | 0.83 (0.17) |
| Statistical Methods for Outcomes (12a) | 0.73 (0.05) | 0.84 (0.05) | 0.78 (0.04) | 1.00 (0.00) |
| Statistical Methods for Other Analyses (12b) | 0.36 (0.16) | 0.28 (0.13) | 0.30 (0.12) | 0.62 (0.20) |
| Participant Flow (13a) | 0.75 (0.09) | 0.79 (0.09) | 0.76 (0.02) | 0.98 (0.03) |
| Participant Loss/Exclusion (13b) | 0.74 (0.07) | 0.69 (0.16) | 0.70 (0.10) | 0.85 (0.14) |
| Periods of Recruitment/Follow-Up (14a) | 0.88 (0.12) | 0.79 (0.15) | 0.82 (0.04) | 0.84 (0.12) |
| Trial Stopping (14b) | 0.65 (0.34) | 0.75 (0.35) | 0.59 (0.25) | 0.73 (0.44) |
| Baseline Data (15) | 0.83 (0.07) | 0.82 (0.14) | 0.81 (0.08) | 0.99 (0.02) |
| Numbers Analyzed (16) | 0.55 (0.24) | 0.43 (0.19) | 0.47 (0.20) | 0.38 (0.11) |

| | | | | |
|---|---|---|---|---|
| Outcome Results (17a) | 0.68 (0.12) | 0.73 (0.04) | 0.70 (0.05) | 0.98(0.03) |
| Binary Outcome Results (17b) | 0.42 (0.15) | 0.41 (0.17) | 0.40 (0.15) | 0.90 (0.04) |
| Ancillary Analyses (18) | 0.51 (0.16) | 0.40 (0.09) | 0.43 (0.07) | 0.83 (0.07) |
| Harms (19) | 0.64 (0.06) | 0.72 (0.04) | 0.68 (0.04) | 0.88 (0.09) |
| Limitations (20) | 0.71 (0.07) | 0.73 (0.02) | 0.72 (0.04) | 0.98 (0.06) |
| Generalizability (21) | 0.58 (0.20) | 0.37 (0.09) | 0.44 (0.10) | 0.71 (0.10) |
| Interpretation (22) | 0.68 (0.05) | 0.70 (0.03) | 0.69 (0.02) | 1.00 (0.00) |
| Registration (23) | 0.91 (0.09) | 0.87 (0.12) | 0.88 (0.08) | 0.96 (0.03) |
| Protocol Access (24) | 1.00 (0.00) | 0.77 (0.33) | 0.83 (0.24) | 0.87 (0.18) |
| Funding (25) | 0.77 (0.10) | 0.83 (0.16) | 0.79 (0.08) | 0.99 (0.02) |
| MICRO | 0.72 (0.02) | 0.71 (0.03) | 0.71 (0.02) | 0.95 (0.01) |
| MACRO | 0.68 (0.04) | 0.63 (0.03) | 0.64 (0.02) | 0.79 (0.03) |

**Table S3:** Item-level results for the fine-tuned BioGPT model.

| **CONSORT Item** | **Precision** | **Recall** | **F1** |
|---|---|---|---|
| Objectives (2b) | 0.85 | 0.85 | 0.85 |
| Trial Design (3a) | 0.80 | 0.50 | 0.62 |
| Changes to Trial Design (3b) | 0.00 | 0.00 | 0.00 |
| Eligibility Criteria (4a) | 0.83 | 0.97 | 0.90 |
| Data Collection Setting (4b) | 1.00 | 0.91 | 0.95 |
| Interventions (5) | 0.88 | 0.75 | 0.81 |
| Outcomes (6a) | 0.78 | 0.96 | 0.86 |
| Changes to Outcomes (6b) | 0.00 | 0.00 | 0.00 |
| Sample Size Determination (7a) | 0.88 | 0.92 | 0.90 |
| Interim Analyses/ Stopping Guidelines (7b) | 0.00 | 0.00 | 0.00 |
| Sequence Generation (8a) | 0.60 | 0.67 | 0.63 |
| Randomization Type (8b) | 0.75 | 1.00 | 0.86 |
| Allocation Concealment (9) | 1.00 | 0.25 | 0.40 |
| Randomization Implementation (10) | 0.54 | 0.70 | 0.61 |
| Blinding (11a) | 0.67 | 0.25 | 0.36 |
| Similarity of Interventions (11b) | 0.00 | 0.00 | 0.00 |
| Statistical Methods for Outcomes (12a) | 0.61 | 0.81 | 0.70 |
| Statistical Methods for Other Analyses (12b) | 0.30 | 0.13 | 0.18 |
| Participant Flow (13a) | 0.69 | 0.67 | 0.68 |
| Participant Loss/Exclusion (13b) | 0.71 | 0.55 | 0.62 |
| Periods of Recruitment/Follow-Up (14a) | 0.70 | 0.64 | 0.67 |
| Trial Stopping (14b) | 0.00 | 0.00 | 0.00 |
| Baseline Data (15) | 0.83 | 0.78 | 0.80 |
| Numbers Analyzed (16) | 0.56 | 0.20 | 0.29 |
| Outcome Results (17a) | 0.72 | 0.69 | 0.70 |
| Binary Outcome Results (17b) | 0.30 | 0.52 | 0.38 |
| Ancillary Analyses (18) | 0.31 | 0.22 | 0.26 |
| Harms (19) | 0.56 | 0.74 | 0.64 |
| Limitations (20) | 0.67 | 0.65 | 0.66 |

| | | | |
|---|---|---|---|
| Generalizability (21) | 0.75 | 0.38 | 0.50 |
| Interpretation (22) | 0.63 | 0.70 | 0.66 |
| Registration (23) | 0.91 | 0.91 | 0.91 |
| Protocol Access (24) | 0.00 | 0.00 | 0.00 |
| Funding (25) | 0.79 | 0.71 | 0.75 |
| MICRO | 0.68 | 0.68 | 0.68 |
| MACRO | 0.59 | 0.55 | 0.55 |

**Table S4:** Item-level results of zero-shot GPT-4 direct inference. Note that these results are obtained on 10 articles.

| CONSORT Item | Precision | Recall | F1 |
|---|---|---|---|
| Objectives (2b) | 0.19 | 0.71 | 0.30 |
| Trial Design (3a) | 0.43 | 0.82 | 0.56 |
| Changes to Trial Design (3b) | 0.00 | 0.00 | 0.00 |
| Eligibility Criteria (4a) | 0.73 | 0.97 | 0.83 |
| Data Collection Setting (4b) | 0.46 | 0.55 | 0.50 |
| Interventions (5) | 0.66 | 0.70 | 0.68 |
| Outcomes (6a) | 0.70 | 0.50 | 0.58 |
| Changes to Outcomes (6b) | 0.00 | 0.00 | 0.00 |
| Sample Size Determination (7a) | 0.73 | 1.00 | 0.85 |
| Interim Analyses/ Stopping Guidelines (7b) | 1.00 | 1.00 | 1.00 |
| Sequence Generation (8a) | 0.64 | 0.90 | 0.75 |
| Randomization Type (8b) | 0.64 | 0.64 | 0.64 |
| Allocation Concealment (9) | 0.64 | 0.88 | 0.74 |
| Randomization Implementation (10) | 0.50 | 0.27 | 0.35 |
| Blinding (11a) | 0.64 | 0.70 | 0.67 |
| Similarity of Interventions (11b) | 0.50 | 0.50 | 0.50 |
| Statistical Methods for Outcomes (12a) | 0.67 | 0.56 | 0.61 |
| Statistical Methods for Other Analyses (12b) | 0.22 | 0.30 | 0.26 |
| Participant Flow (13a) | 0.53 | 0.83 | 0.65 |
| Participant Loss/Exclusion (13b) | 0.38 | 0.17 | 0.23 |
| Periods of Recruitment/Follow-Up (14a) | 0.60 | 0.5 | 0.55 |
| Trial Stopping (14b) | 0.00 | 0.00 | 0.00 |
| Baseline Data (15) | 0.90 | 0.77 | 0.83 |
| Numbers Analyzed (16) | 0.35 | 0.31 | 0.33 |
| Outcome Results (17a) | 0.40 | 0.41 | 0.41 |
| Binary Outcome Results (17b) | 0.00 | 0.00 | 0.00 |
| Ancillary Analyses (18) | 0.32 | 0.32 | 0.32 |
| Harms (19) | 0.25 | 0.95 | 0.40 |
| Limitations (20) | 0.36 | 0.48 | 0.41 |
| Generalizability (21) | 0.29 | 0.71 | 0.42 |
| Interpretation (22) | 0.52 | 0.42 | 0.47 |
| Registration (23) | 1.00 | 0.83 | 0.91 |
| Protocol Access (24) | 0.07 | 1.00 | 0.13 |

| | | | |
|---|---|---|---|
| Funding (25) | 0.71 | 0.96 | 0.82 |
| MICRO | 0.48 | 0.54 | 0.51 |
| MACRO | 0.47 | 0.58 | 0.49 |

### 5. Data augmentation examples

**Table S5:** Data augmentation outcomes using different methods. The original sentence used for GPT-4 paraphrasing, EDA, and UMLS-EDA is *The packaging and labeling of the study drug kits were based on a separate drug packaging randomization schedule*, with the label Allocation Concealment Mechanism (9). The item description used for generative augmentation is "the mechanism used to implement the random allocation sequence, describing any steps taken to conceal the sequence until interventions were assigned, with specifics". EDA augmentation randomly inserts a token "randomisation" into the sentence. UMLS-EDA introduces "[mental process]" into the sample.

| Augmentation method | Augmentation result |
|---|---|
| GPT-4 generative | *To achieve random allocation, we used a shuffled deck of cards each representing a treatment category, keeping every card faced down until the time came to assign treatment legitimately.* |
| GPT-4 rephrasing | *A distinct randomization schedule for drug packaging was utilized for the arrangement and tagging of the study drug kits.* |
| EDA | *The packaging and labeling of the study drug randomisation kits were based on a separate drug packaging randomization schedule.* |
| UMLS-EDA | *The packaging and labeling [mental process] of the study medication kits were based on a separate drugs packaged randomization schedule.* |

### 6. Error analysis

We analyzed the errors made by the best-performing PubMedBERT model. Most error cases involved sentences predicted to report a CONSORT item different from the original label (37.2% of errors). For some of these cases, the true label was among the predictions, but additional labels were predicted (8.3%). Similarly, for 6.3% of the errors, at least one true label was correctly predicted, but some other labels were missed. In 1.1% of the cases, there was at least one label overlap, while some labels were missed and others were incorrectly predicted. These types of overlaps, accounting for 15.7% of the total number of errors could be considered less fatal. About 29.3% of the errors involved negative sentences that were predicted to report a CONSORT item, and the rest (33.5%) were those labeled with a CONSORT item but for which no CONSORT label was predicted. We provide the samples for these error types in Table S5.

**Table S6:** Samples for different error types. The labels shown are Outcomes (6a), Outcome Results (17a), Binary Outcome Results (17b), and Ancillary Analyses (18).

| **Error Type** | **Sentence** | **True** | **Predicted** |
|---|---|---|---|
| Predicted CONSORT item different from the true label | *The findings from the adjusted analysis were similar to those of the unadjusted analysis (odds ratio 1.17 (0.62 to 2.21), P=0.62).* | [‘17b’] | [‘17a’] |
| Some CONSORT items are missed. | *Patients were directed to fix the adhesive ring with the hole over the verruca and to squeeze a little ointment into the hole and directly on to the verruca.* | [‘11a’, ‘6a’] | [‘6a’] |
| Additional CONSORT items are predicted. | *The number of plantar warts at the start of the study was not found to be an important predictor of outcome (odds ratio 0.81 (0.65 to 1.01)).* | [‘17a’] | [‘17a’, ‘18’] |

The confusion matrix for the best-performing PubMedBERT model is in Figure S1. The most confusion occurred between Outcome Results (17a) and related items Binary Outcome Results (17b) and Ancillary Analyses (18). This was followed by Statistical Methods for Outcomes (12a) and Statistical Methods for Other Analyses (12b). We note that inter-annotator agreement for these labels was lower, as annotators often confused them as well[2]. For practical use, it might be practical to merge these labels. The label that was most often completely missed or incorrectly predicted was Interpretation (22), which is a broad and diffuse category, similar to Background (2a), and its utility might be considered debatable.

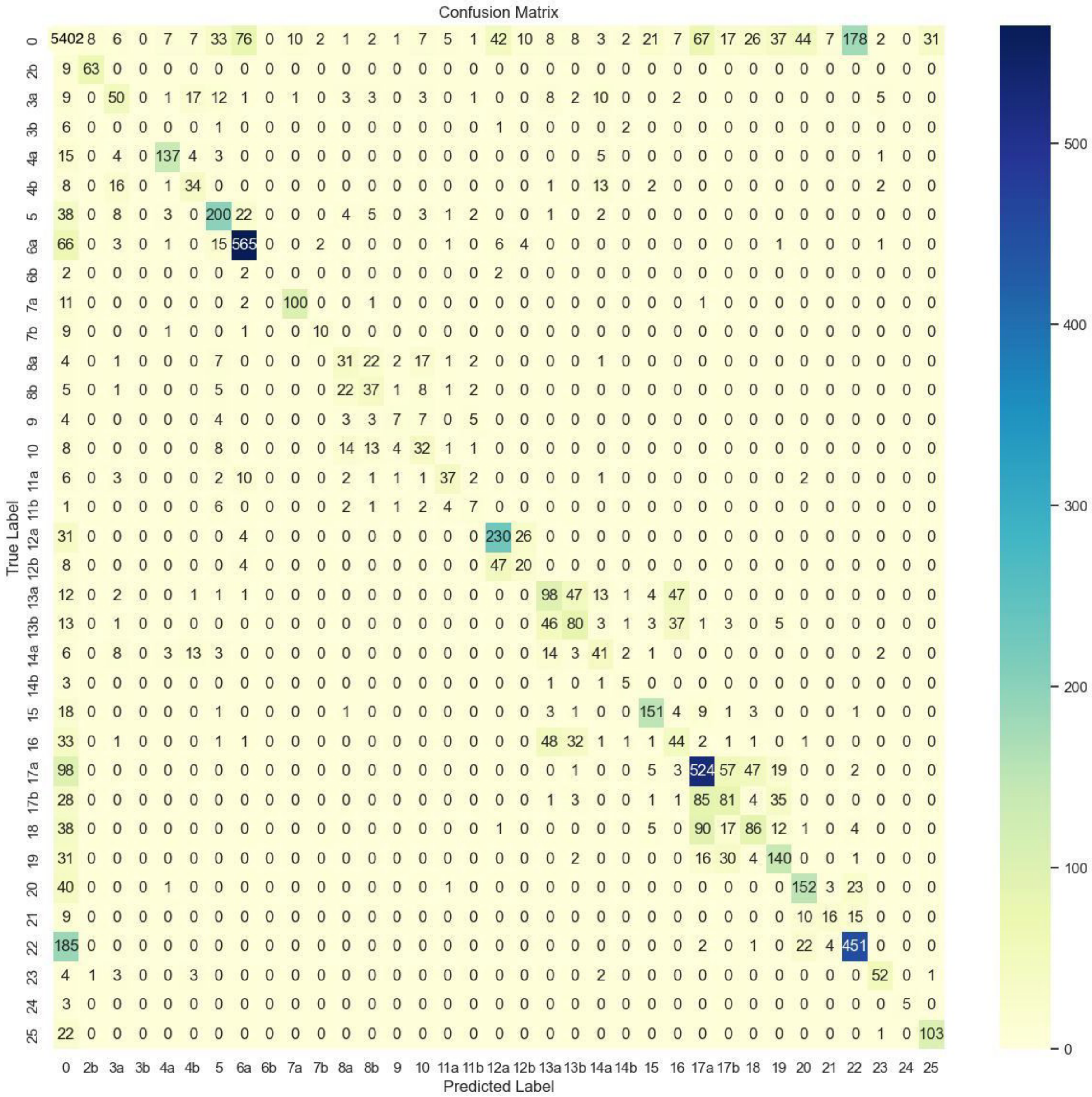


**Figure S1:** Confusion matrix for the best-performing model.
